# Multi-modal data to identify key factors influencing lung injury in ARDS patients undergoing invasive mechanical ventilation: A prospective observational study protocol

**DOI:** 10.1101/2025.09.09.25335458

**Authors:** Zhimei Duan, Di Lian, Kaifei Wang, Ye Hu, Han Fu, Ruoxuan Wen, Ying Zhao, Xingshuo Hu, Pan Pan, Jianqiao Xu, Jin Chen, Li Xiao, Lin Wang, Xiao Yu, Xiaobo Han, Wuxiang Xie, Fei Xie, Lixin Xie, Zhihai Han

## Abstract

**Background:** Patients with moderate to severe acute respiratory distress syndrome (ARDS) exhibit extremely poor prognoses following mechanical ventilation, with mortality rates as high as 40% to 55%. Despite extensive research into ARDS classification and prognostic assessment, the disease’s pathogenesis remains incompletely understood, and there remains a critical lack of specific biomarkers and effective therapeutic targets for its prevention and management. The core challenges lie in two key areas: First, ARDS demonstrates marked heterogeneity in etiology, pathophysiology, and pathogenesis. Second, current definitions of ARDS phenotypes are often confined to clinical symptoms and routine diagnostic indices, lacking integrated analysis of deeper mechanistic indicators, thereby limiting the stability and clinical utility of existing classification systems.

**Methods/Design:** We designed a prospective multicenter cohort study incorporating multi-omics analyses. The study plans to enroll over 200 patients with moderate to severe ARDS receiving mechanical ventilation across 10 medical centers. Peripheral blood and bronchoalveolar lavage fluid (BALF) samples will be collected on the first 24 hours after enrollment and at extubation for metagenomic/meta-transcriptomic sequencing, bulked RNA sequencing, single-cell transcriptomics, proteomics, and metabolomics analyses. Concurrently, comprehensive monitoring of physiological indices, electrical impedance tomography, transpulmonary pressure, pulmonary ultrasound findings, and other relevant parameters will be conducted during the enrollment. Study participants will be stratified by survival and mortality outcomes to analyze the dynamic trends of all measured indices and their underlying molecular mechanisms.

**Discussion:** Through comparative analysis of multi-omics data, we aim to identify specific markers and risk factors associated with distinct clinical trajectories of ARDS, further clarifying the key determinants of lung injury. Ultimately, this research will reveal critical immune cell subtypes that govern ARDS onset and prognosis, offering novel insights and therapeutic targets to advance precision medicine for ARDS.

**Trial registration:** ClinicalTrials.gov NCT 05922826. Registered on 27 Six 2023.

## Introduction

Acute Respiratory Distress Syndrome (ARDS) is a complex clinical syndrome triggered by various risk factors, including pneumonia, non-pulmonary infection, trauma, blood transfusion, burns, aspiration, and shock. These factors can increase the permeability of the pulmonary vascular endothelium and alveolar epithelium, leading to pulmonary edema and gravity-dependent atelectasis, which form the pathological basis of ARDS. Characterized by bilateral pulmonary infiltrates and non-cardiogenic pulmonary edema, ARDS is marked by progressive respiratory distress and refractory arterial hypoxia[1–4]. Due to the high degree of clinical and biological heterogeneity among ARDS patients, it remains challenging to develop fundamentally effective treatments to reduce mortality, with in-hospital mortality rates remaining as high as 40%-55% worldwide[5, 6]. Currently, the only clinically proven effective treatments are primarily supportive, including lung-protective mechanical ventilation, prone positioning, and high positive end-expiratory pressure (PEEP). In the era of precision medicine, integrating various forms of information (e.g. clinical features, imaging, physiology, biological tests, and multi-omics analyses) to accurately subtype ARDS could enable the pairing of treatments with treatable characteristics. This approach may more reliably predict patients’ responses to therapies and interventions, representing a future research direction for ARDS[2, 7, 8].

The study of mechanical ventilation has long been the focus of clinical research in ARDS. Currently, various methods are employed to evaluate the therapeutic efficacy and prognosis of these patients. Electrical impedance tomography (EIT) has been applied to assess the degree of pulmonary collapse in gravity-dependent regions in patients with COVID-19 secondary to ARDS. It was found that the degree of collapse in gravity-dependent areas in supine position is significantly associated with the improvements in oxygenation during prone positioning, enabling accurate prediction of the therapeutic effect of prone position ventilation[9]. Transpulmonary pressure is widely recognized as one of the methods for titrating individualized PEEP in patients with ARDS, which can avoid lung injury caused by excessive stress and strain, alleviate clinical symptoms, predict the clinical outcome, and improve the survival rate of patients[10]. However, transpulmonary pressure-guided PEEP titration is based on a single-compartment model, which has its limitations. For patients with high heterogeneity, such as those with ARDS, the method of carrying out individualized PEEP titration remains controversial. Critical care ultrasound can monitor the changes in lung ventilation in ARDS patients in real-time. By using the Lung Ultrasound Score (LUS), it dynamically monitors the changes in lung ventilation during the lung recruitment process, facilitating the individualized titration of mechanical ventilation parameters. Additionally, it can monitor the effects of prone positioning and predict the clinical outcomes in ARDS patients[11–13].

In recent years, the rapid development of molecular biology and bioinformatics has gradually deepened our understanding of the pathophysiology of ARDS. Recognizing the heterogeneity of ARDS, researchers have used muti-omics technologies, such as genomics, transcriptomics, proteomics, metabolomics and metagenomics, to identify targeted therapies for different ARDS subgroups[14–17]. In a study using k-means cluster analysis of whole blood microarray data, three different subtypes, called CHOP (Children’s Hospital of Philadelphia) ARDS transcriptome subtypes (CATS), were identified in 96 pediatric ARDS patients. CATS1 was mainly associated with adaptive immunity and T cell pathway, CATS2 with complement pathway, and CATS3 with G protein-coupled receptor signaling pathway. Patients with CATS1 subtype exhibited the lowest survival rate, corresponding to a 32% mortality rate, whereas those with the CATS3 subtype showed the highest, with an 8% mortality rate[18]. This stratification is highly beneficial for clinical diagnosis and significantly reduces the use of unnecessary medications.

A study on mouse transcriptomes confirmed that the increased gene and protein expression of *ELAVL-1/HUR* and *GSK3β* is related to the poor therapeutic outcomes in ARDS[19]. Additionally, high expression levels of p300/CBP in peripheral blood have been identified as a risk factor for 28-day mortality in ARDS[20]. Single nucleotide polymorphisms (SNP) of mRNA, including *TNF-α* (*rs1800629)*, *IL-6 (rs1800796)*, and *MyD88 (rs7744)* are markers of increased risk and poor prognosis in ARDS[21]. The gene expression patterns of alveolar macrophages (AMs) and Peripheral Blood Mononuclear Cells (PBMCs) in ARDS patients are highly heterogeneous. It has been suggested that the up-regulated expression of pro-inflammatory genes in PBMCs is associated with worse clinical outcomes[22]. The down-regulation of *METTL16*, *FTO*, *METTL3*, *KIAA1429*, *RBM15*, *ALKBH5*, *YTHDF2*, *YTHDF3*, *YTHDC2*, and *IGFBP2* is related to lipopolysaccharide induced M6A methylation and is involved in regulating inflammation and the development of lung injury[23].

A proteomic study on ARDS sub-phenotype using BALF from the survivors and non-survivors of ARDS in early (<7 days) and late (>8 days) stages revealed that, the activation of immune response, wound healing, and multiple pathways involved in blood coagulation were more prominent in the survivors, while collagen synthesis and carbohydrate catabolism were more abundant in the non-survivors[24]. Viswan et al. have identified distinct biological endotypes of ARDS[25]. They found specific amino acids such as isoleucine, leucine, lysine / arginine, tyrosine, and threonine in BALF, and proline, glutamic acid, and phenylalanine in serum. The presence of these metabolites and their association with SOFA and APACHE II scores provide a robust predictor of mortality in ARDS patients. Furthermore, a comparison of metabolomics characteristics between ARDS patients and healthy controls revealed higher levels of phenylalanine, D-phenylalanine, and phenylacetylglutamine in non-survivors[26]. Shortt et al. identified a novel SNP associated with ARDS through whole exons sequencing (WES), which could be a potential new biomarker for ARDS. Although gene regulation can alleviate some symptoms of ARDS, no single gene or gene combination specific to ARDS has been found in experimental animal or human studies[27].

Metagenomic sequencing of BALF samples from patients with ARDS showed significant diversity changes in the microecology of the lower respiratory tract. In a healthy state, the microbiota of the human respiratory tract remains relatively stable, with bacteria maintaining a symbiotic relationship that supports normal respiratory function[28, 29]. However, during ARDS, the influence of inflammatory reaction, immune imbalance and other factors disrupt this equilibrium. Some beneficial bacteria may decrease, while pathogenic bacteria or opportunistic pathogenic bacteria increase. These changes of microecological diversity have been found high relations to the severity, treatment response, and prognosis of ARDS[30, 31].

Despite extensive research on ARDS typing, accurate treatment and poor prognosis, the disease progression of ARDS remains unclear, and there are still no specific biomarkers and effective targets for prevention and treatment for ARDS. This study will conduct a multicenter data collection, including clinical indicators, mechanical ventilation parameters (such as PEEP titration data), lung ultrasound findings, EIT metrics, and other relevant parameters, from mechanically ventilated patients with moderate to severe ARDS. Concurrently, the study will integrate meta-genomic and meta-transcriptomic next-generation sequencing, bulked RNA sequencing (RNA-seq), single-cell RNA sequencing (scRNA-seq), proteomic and metabolomic data from the patients’ blood and bronchoalveolar lavage fluid (BALF).

By summarizing changes in respiratory therapy-related parameters and exploring quantitative indicators for evaluating lung injury, the study aims to identify key factors that determine the development and prognosis of ARDS, thereby providing insights for its precision treatment.

## Materials and Methods

### Purpose of the study

The study aims to identify risk factors associated with differential survival and mortality outcomes in mechanically ventilated patients with moderate or severe ARDS who exhibit varying prognoses. This objective will be accomplished by investigating the differences and dynamic changes in clinical examinations, respiratory mechanics, lung ultrasound findings, and multi-omics analyses, as well as by further exploring their associations with prognosis. Additionally, the study seeks to screen for functionally significant genes and candidate pathways, with the goal of identifying key immune cell subtypes that determine the onset and prognosis of ARDS, providing novel insights and therapeutic targets to advance the precision treatment of ARDS.

### Study design

In this study, a prospective multicenter clinical cohort study will be conducted to observe the changes of lung injury, EIT, transpulmonary pressure, lung ultrasound, and other indexes in patients with ARDS. The effects on lung injury in patients with ARDS from the perspectives of respiratory mechanics, ventilation and perfusion, and lung imaging will be evaluated. The key indicators to provide clinical data support for the establishment of a 28-day death prediction model for patients with ARDS will be screened. The detailed technical route is shown in the following **fig 1**. The specific sampling strategies and multi-omics detections were illustrated in **fig 2**.

**Fig 1.**
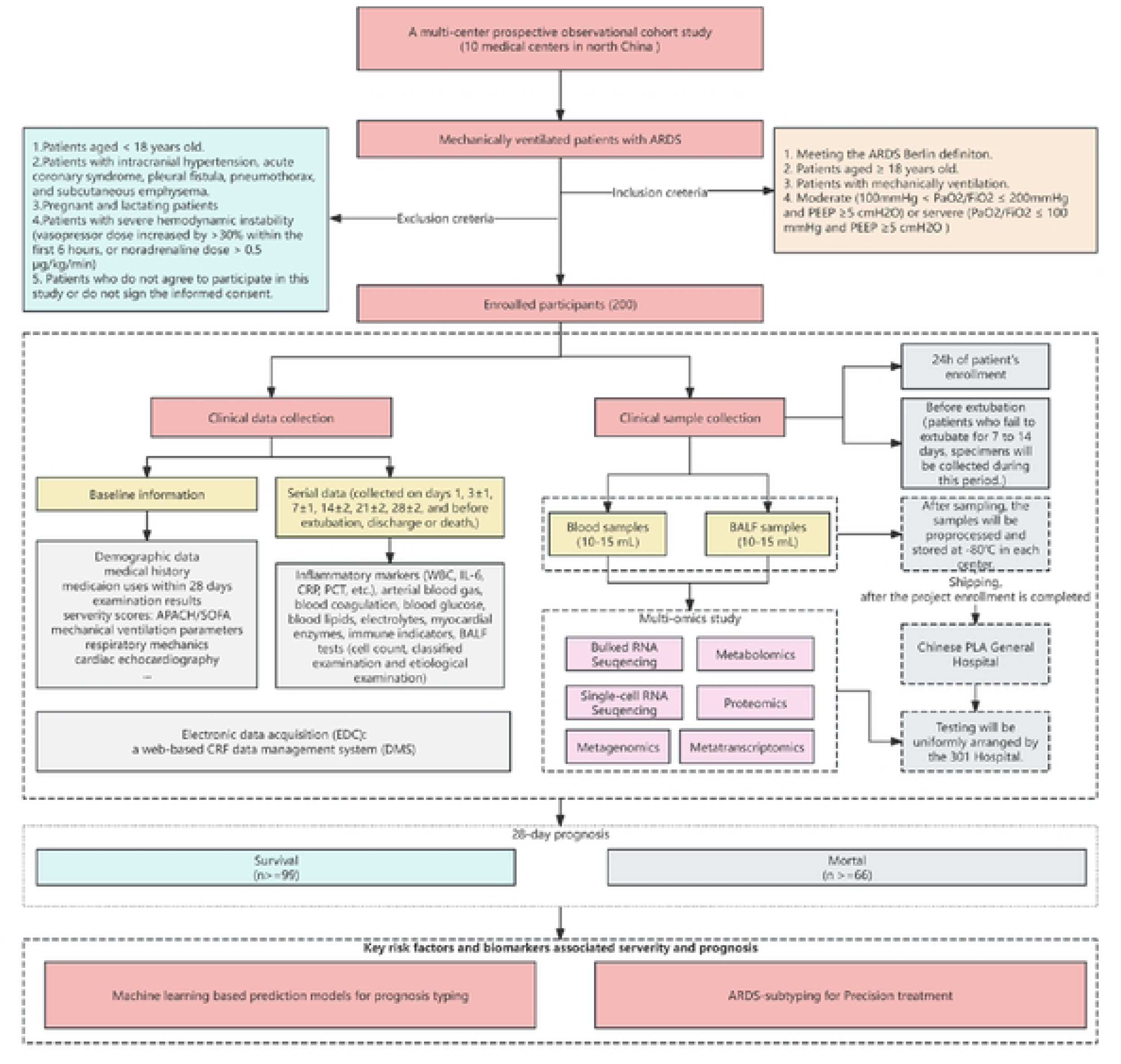
Flowchart showing the study design. After the patient meets the inclusion and exclusion criteria and provides informed consent, baseline data and clinical samples are collected. Blood and BALF samples are obtained on the first day of enrollment and prior to extubation. During this period, all clinical test results, monitoring data, and treatment records are gathered, and the clinical outcomes at 28 days are documented. Multi-omics analyses, including transcriptomics, proteomics, metabolomics, and genomics, are performed on the collected samples. Clinical data are further analyzed using stratification techniques. All data are integrated, and machine learning approaches are employed to identify key factors contributing to lung injury in ARDS patients undergoing invasive mechanical ventilation.

**Fig 2.**
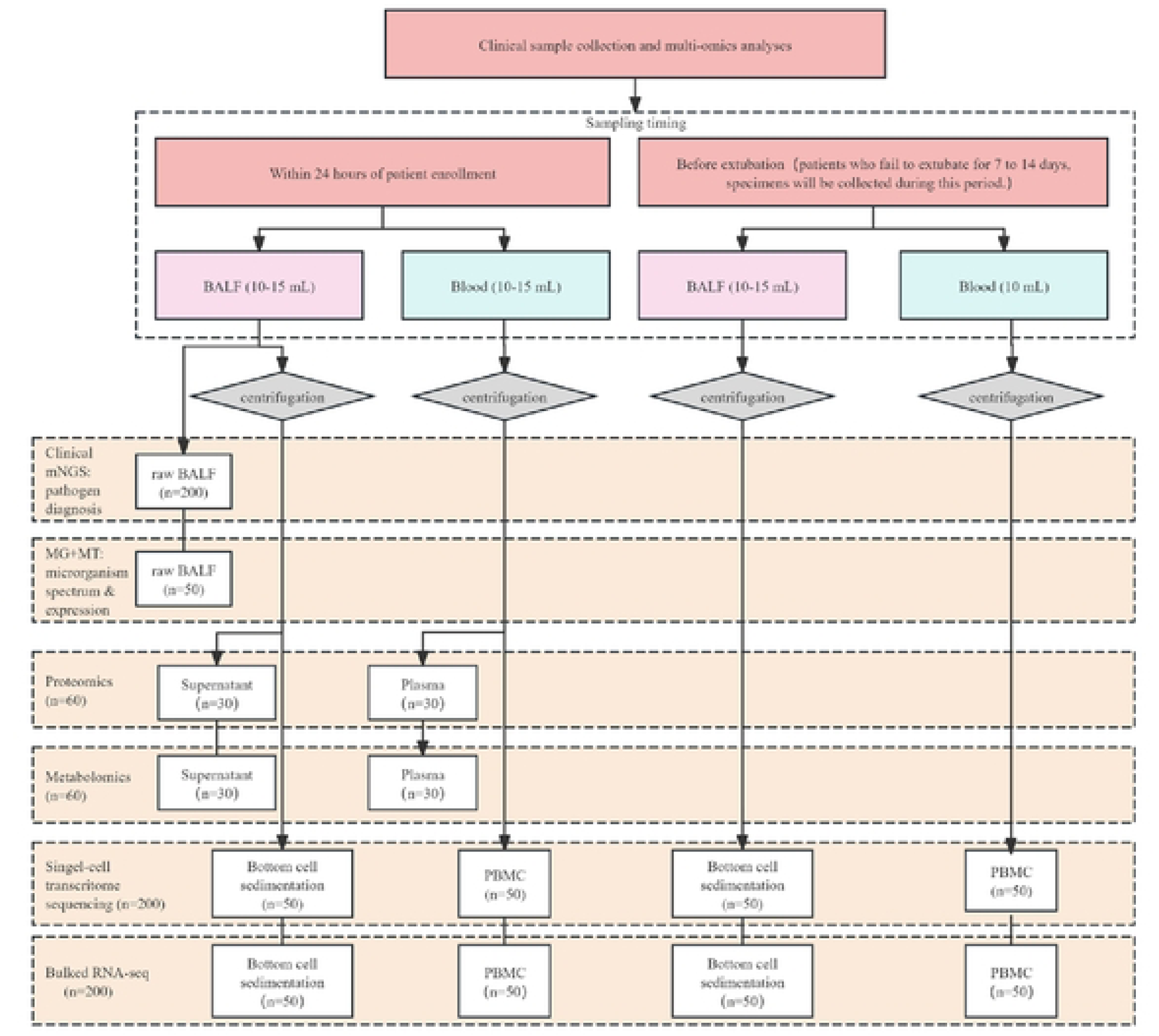
Sample collection, pretreatment, and multi-omics detection process. A total of six-type omics assays will be performed, with sample utilization stratified by type and collection time point: BALF samples collected within 24 hours of enrollment will first undergo clinical mNGS for pathogen diagnosis, while the original fluid will also be used for metagenomic microecology analysis and meta-transcriptomic microbial expression detection; after centrifugation, the supernatant will be subjected to proteomic and metabolomic assays, and the bottom cell sedimentations will be used for transcriptomic and single-cell transcriptomic analyses. Concurrently collected blood samples, following centrifugation, will have plasma used for proteomic and metabolomic assays, PBMCs utilized for transcriptomic and single-cell transcriptomic analyses. For BALF samples collected prior to extubation, centrifugation-derived bottom cell sedimentations will be used for transcriptomic and single-cell transcriptomic analyses; blood samples collected at this time, after centrifugation, will have PBMCs used for transcriptomic and single-cell transcriptomic analyses.

### Participant recruitment

Participants will be recruited from ten medical centers in tertiary general hospitals in North China, including the First Medical Center of Chinese PLA General Hospital, the Sixth Medical Center of Chinese PLA General Hospital, the Eighth Medical Center of Chinese PLA General Hospital, Beijing Shijitan Hospital, Beijing Anzhen Hospital, AMHT Group Aerospace 731 Hospital, Zhengzhou Central Hospital, Henan Provincial People’s Hospital , the First Affiliated Hospital of Zhengzhou University and First Hospital of Shanxi Medical University.

Patients with ARDS will be invited to participate in this study upon admitting the branch center. Those who meet the inclusion criteria and provide signed informed consent will enter the screening period (**Supplementary materials: additional file 1**). During the screening visit, trained researchers (physicians) will confirm the eligibility based on symptoms, laboratory results, and chest X-ray or CT image, and review every inclusion and exclusion criteria. Patients who meet the exclusion criteria will be excluded from this study within 24 hours. The recruitment period is set for 24 months, spanning from November 2023 to November 2025.

### Inclusion and exclusion criteria

In this trial, patients with moderate or severe ARDS and experienced mechanical ventilation will be included for inclusion selection. The definition of ARDS refers to the Berlin definition of ARDS[3]. Patients with moderate ARDS are defined as those with 100 mmHg < PaO_2_/FiO_2_ ≤ 200 mmHg with PEEP ≥5 cmH_2_O, and severe ARDS refers to those with PaO_2_/FiO_2_ ≤ 100 mmHg with PEEP ≥5 cmH_2_O.

Patients meeting any of the following criteria will be excluded from the trial:

1. Patients aged less than 18 years old.
2. Patients with intracranial hypertension, acute coronary syndrome, pleural fistula, pneumothorax, and subcutaneous emphysema.
3. Pregnant and lactating patients.
4. Patients with severe hemodynamic instability (vasopressor dose increased by >30% within the first 6 hours, or noradrenaline dose > 0.5 μg/kg/min).
5. Patients who do not agree to participate in this study and patients do not sign the informed consent.

### Data collection

The formal baseline data and serial data of the participants will be collected. Three case report forms (CRFs) have been designed for different stages of patient enrollment management (**Supplementary materials: additional file 2-4**).

The formal baseline data includes the following aspects:

Demographic data: hospitalization number, name, sex, age, main cause of admission, ethnicity, smoking history, and body mass index.

Medical history: hypertension, coronary heart disease, diabetes, kidney disease, hepatopathy, cerebrovascular disease, autoimmunity disease, hemopathy, pulmonary disease, and cancer.

Medication use within 28 days: antibiotics, hormones and immunosuppressants, vasoactive agent, neuromuscular blockers, sedative and analgesic drugs.

Physical examination findings: blood pressure, pulse rate, heart rate, respiratory rate, and temperature.

Severity scores: the Acute Physiology and Chronic Health Evaluation (APACHE) II and the Sequential Organ Failure Assessment (SOFA) score.

Mechanical ventilation parameters (on day 1 and before extubation): ventilation mode, inhaled oxygen fraction, PEEP, frequency, tidal volume, flow rate, expiratory tidal volume, peak pressure, plateau pressure, airway resistance.

Respiratory mechanics (on day 1 and before extubation): the respiratory system compliance (C_rs_), static compliance of the lungs (C_lung_), respiratory system airway resistance (R_rs_), esophageal pressure (P_es_), transpulmonary pressure (P_tp_), transpulmonary driving pressure (△P_tp_), driving pressure (△P), dead space fraction (V_D_/V_t_) and mechanical power (MP).

Cardiac echocardiography (on day 1 and before extubation): measurement of left ventricular ejection fraction.

Pulmonary ultrasound score and pulse indicator continuous cardiac output parameters.

Serial data: The serial data will be collected on days 1, 3±1, 7±1, 14±2, 21±2, 28±2, and before extubation, discharge or death, including inflammatory markers (white blood cell count, neutrophil count and proportion, interleukin-6, C-reactive protein, procalcitonin, etc.), arterial blood gas, coagulation, blood glucose, blood lipids, electrolytes, myocardial enzymes, immune indicators, BALF tests (cell count, classified examination and etiological examination).

### Data management

In this study, we employ a web-based CRF data management system (DMS) for data collection and centralized management throughout the experiment. After new participants are enrolled, data collectors will visit sub-centers weekly to input required information into the online DMS (https://h6world.cn). The data administrator is responsible for collecting data monitored by independent external supervisors. Questions or outliers should be answered by the quality control officer. 10% of the subjects will be randomly selected and tested with the original questionnaire to evaluate the bit error rate. Based on the bit error rate, the subsequent monitoring plan will be arranged. If an error occurs, the incorrect data in the database must be corrected and all modifications must be documented in the electronic data collection system. Once data entry and resolution of issues are complete, the research data will be locked by the lead researcher, preventing any further alterations by the researchers. No compensation will be provided.

### Sample collection, and multi-omics profiling

For enrolled patients, peripheral blood and BALF samples will be collected at two time points: within 24h after enrollment and before tracheal extubation. For patients who cannot be extubated within 7–14 days, samples will be collected during this 7-14 day window (**Fig 2**).

For blood sampling, patients must be in a fasting state prior to blood collection. A total of 10-15 mL of blood will be collected using anticoagulant tubes, which will then undergo pretreatment before separate storage or detection. Within four hours of sampling, centrifugation will be conducted at 3000 r/min for 10 minutes. The resulting plasma will be aliquoted into 1.5 mL EP tubes at 500 μL per tube for subsequent proteomics and metabolites detection. For the remaining sample, equal volumes of PBS and Ficoll lymphocyte separation solution will be added, followed by centrifugation at 1000g for 20 minutes to collect PBMCs. After adding 5 mL of PBS, the mixture will be centrifuged at 400g for 5 minutes, the supernatant will be discarded, and PBMCs will be resuspended in cell cryopreservation solution before aliquoting into EP tubes at 1 mL per tube. The PBMC samples will be used for RNA-seq and scRNA-seq. The samples will be stored at −80℃ before multi-omics detection.

For BALF sampling, two tubes of BALF samples will be collected. One tube, containing no less than 10 mL, will undergo pretreatment for subsequent RNA-seq, scRNA-seq, proteomics and metabolites detection. The other tube, with a volume of at least 5 mL, will be directly submitted for clinical mNGS pathogen testing, as well as for metagenomic microbial spectrum analysis and meta-transcriptomic expression profiling. For reserved samples, 6 mL of BALF will be centrifuged at 4000g for 10 minutes.

The supernatant will then be aliquoted into 1.5 mL EP tubes at 400 μL per tube, while 1 mL of Trizol will be added to the remaining cell pellet, followed by vortex mixing. All aforementioned samples, along with the remaining BALF, will be labeled and stored at −80 ℃ . The supernatant will be used for proteomics and metabolites detection, while the bottom cell-sedimentation will be used for RNA-seq and scRNA-seq.

Upon completion of participant enrollment, all stored samples will be uniformly transported via cold chain to the Chinese PLA General Hospital for subsequent multi-omics testing.

### Follow–up schedules

At the 28-day mark, patients will be assessed for the primary outcome of survival or death. At this point, the attending physician will report the primary outcome, and the data collection database will then document the 28-day mortality rate, as well as the duration of ICU and hospital stays, along with other relevant outcomes. During the trial period, specific measurements will be taken at predetermined time points: day 1, day 3±1, day 7±1, day 14±2, day 21±2, day 28±2, and before extubation, discharge or death. The following parameters will be recorded: inflammatory markers, arterial blood gas, coagulation, blood glucose, blood lipids, electrolytes, myocardial enzymes, immune indexes, cell count and classification of BALF and etiological examination. 28-day administration of antibiotics, neuromuscular blockers, vasoactive agent, hormones and immunosuppressants will be tracked. The measurement and collection schedule of this experiment are summarized in the follow-up form (**Table 1**).

**Table 1.**
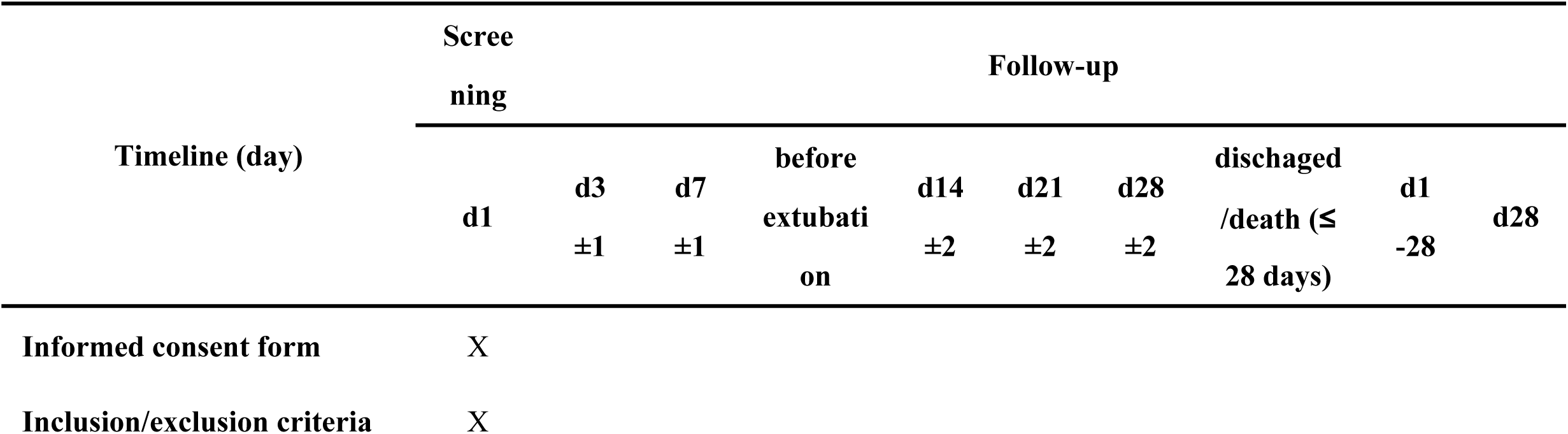
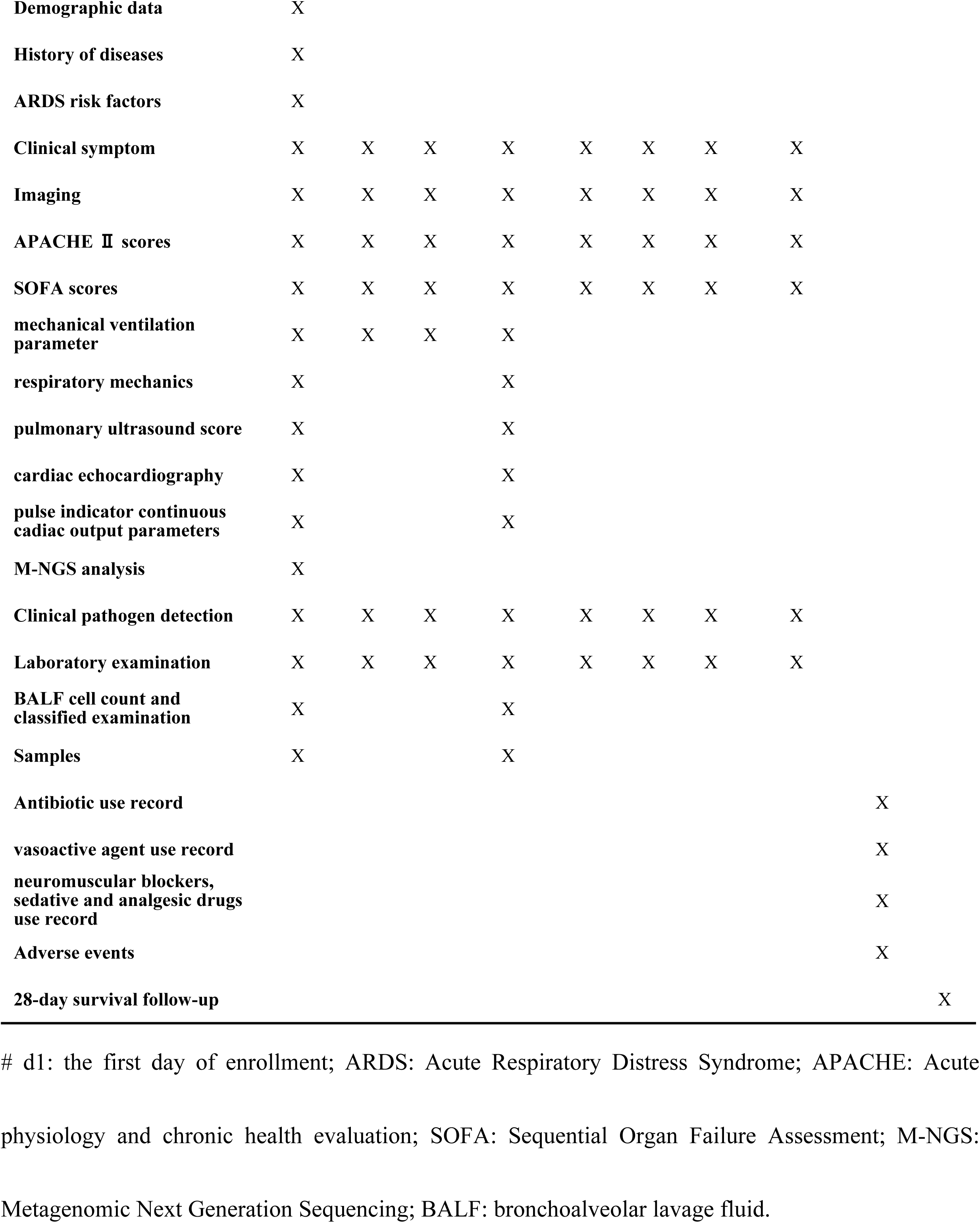
The schedule of measurements and visits of this trial.

### Outcomes

The primary outcome of this study is the 28-day clinical outcome, which includes survival and death. The secondary outcomes include demographic data, clinical data, 28 days of ICU hospitalization and length of stay. For patients with moderate to severe ARDS undergoing invasive mechanical ventilation, additional outcomes include Bulk RNA Sequencing, and Single-Cell RNA Sequencing analyses of peripheral blood and BALF will be collected on 24h of inclusion and before extubation. For patients unable to be extubated within 7 to 14 days, samples will be collected during this period. Respiratory mechanics parameters, severe ultrasound indexes, EIT data, the correlation of physiological parameters will also be evaluated using binary analysis.

### Sample size calculation

The study hypothesizes that the developed model will achieve an area under the ROC curve of at least 0.85 for predicting the 28-day mortality in ARDS patients, and the goal is to test if this value is greater than 0.70, which is considered the threshold for good discriminant ability. The study is designed with α = 0.05 and β = 0.10, and the proportion of lost follow-up or withdrawal is less than 10%. The sample size was calculated using PASS-16 software, which determined that at least 66 deaths must be observed for the study. Referring to previous literature, the 28-day mortality rate of ARDS patients is about 40%, and it is estimated that 165 patients with ARDS need to be enrolled in the study.

### Statistical analysis

For continuous variables with a normal distribution, the t-test or analysis of variance will be employed to compare means. For those with a non-normal distribution, the rank sum test will be used to compare medians. A Cox regression model will be constructed to establish a predictive model for 28-day mortality in patients with ARDS. The predictive performance of the model will be evaluated using the area under the receiver operating characteristic (ROC) curve, as well as sensitivity and specificity.

### Ethics and dissemination

This trial adheres to the Declaration of Helsinki and the guidelines for the quality management of drug clinical trials. Prior to enrollment, each subject or their legal representative will receive a comprehensive explanation of the study and will be required to sign the informed consent form (**Supplementary materials: additional file 1**). Subject data stored in the DMS will be password-protected, with access restricted exclusively to authorized users at the appropriate permission level to ensure the security of data used for statistical analysis. This study has been approved by the Ethics Committee of the PLA General Hospital (Approval Number: 2022113030901836).

### Quality control

Prior to the start of the study, key researchers have received training on the Drug Clinical trial quality Management Code (GCP). All researchers underwent comprehensive training to ensure trial quality, including research protocols, informed consent forms, case reports, standard operating procedures for subject data collection, and methods for biological sample collection and preservation. In addition, a detailed researcher’s manual was developed to ensure compliance with the program. During the study, biological samples will be tested by trained professionals and genetic testing companies.

An executive committee, comprising members of the expert group, including expert committees composed of clinicians, statisticians, and quality managers, will oversee clinical trials progress, organize trial-related seminars, and manage data collection and analysis. Quality control meetings will be convened as needed.

A dedicated quality control team, composed of qualified inspectors, will conduct research audits and monitoring every two weeks. Inspections will cover the completeness of study documentation and informed consent records, adherence to inclusion/exclusion criteria, accuracy of raw data, management of adverse events (AEs) and serious adverse events (SAEs), sample storage conditions, and data collection records. Findings will be reported to key researchers and relevant committees. Adverse events, such as cardiac events, drug side effects or deaths, will be recorded using data collected for secondary outcomes, such as mortality and hospitalization. Participating facilities must immediately report all AEs to the quality control department. Serious events will be evaluated by the principal researcher or other clinical members of the trial management team to determine the cause and prognosis. All incidents will be tracked until resolution or decision to discontinue follow-up, with relevant information shared among researchers.

Furthermore, any modifications to the protocol, including changes to study type, design, subject population, sample size or pilot procedure, must undergo formal revision and approved by the Ethics Committee. Updates will be documented in Clinical Trials registry. If a protocol change may affect trial-related risks, the informed consent of the recipient will be re-obtained.

### Patient and public involvement

No patients or members of the public were involved in the design, conduct, reporting, or dissemination of this research study.

## Discussion

ARDS remains a critical challenge in critical care medicine, characterized by acute onset triggered by diverse pulmonary and extrapulmonary insults. Pathologically, it is defined by alveolar epithelial and pulmonary microvascular endothelial injury, culminating in widespread pulmonary inflammation. As a complex clinical syndrome, ARDS exhibits striking heterogeneity across its etiology, pathophysiology, clinical presentation, and imaging characteristics, which has hindered the development of universally effective therapies[2, 3, 7]. While recent advancements in medical and biological technologies have deepened our understanding of this heterogeneity, significant gaps persist in translating such insights into improved patient outcomes[32].

Against this backdrop, the paradigm of precision medicine has emerged as a transformative approach, emphasizing the need to tailor diagnosis and treatment to individual patient characteristics[33, 34]. This shift has underscored the value of integrating advanced technologies, including molecular biology, bioinformatics, scRNA-seq, transcriptomics, microbiome analysis, proteomics, and metabolomics, in ARDS research. These tools hold promise for unraveling dynamic changes in key molecules, signaling pathways, and cell subsets during ARDS initiation and progression, ultimately enabling individualized and targeted therapeutic strategies. Such efforts represent the forefront of future ARDS research, aiming to overcome the limitations of one-size-fits-all approaches.

The present study contributes to this evolving landscape through several key strengths. It is the first large-scale, multicenter observational cohort study with rigorous quality control focused on mechanically ventilated patients with moderate to severe ARDS. By systematically collecting comprehensive baseline and clinical data, alongside BALF and peripheral blood samples at two critical time points (day 1 post-admission and pre-extubation), the study enables in-depth multi-omics analyses, including bulked and single cell transcriptomics, microbiome, proteomics and metabolomics. These data will address two pivotal questions: (1) the predictive value of clinical and respiratory mechanics indices for 28-day mortality in this high-risk population, and (2) the interplay between lower respiratory tract microecology (encompassing bacteria, viruses, fungi, etc.) and host immunity across differing clinical outcomes. Together, these investigations aim to provide novel insights into precision treatment strategies for mechanically ventilated patients with moderate to severe ARDS.

Despite its comprehensiveness, the study has inherent limitations. First, while the sample size is substantial, its representativeness of the broader ARDS population remains to be verified; further validation with an expanded cohort may be necessary to confirm the robustness of the findings. Second, the multicenter design, while strengthening generalizability, introduces challenges related to ensuring consistency in data collection, sample processing, and outcome assessment across sites, which must be rigorously addressed to maintain result reliability.

Looking forward, longitudinal studies are warranted to evaluate long-term outcomes of ARDS survivors and assess the durability of any predictive models derived from the current data. Additionally, interventional trials are critical to translating the study’s findings into clinical practice, testing whether targeting identified molecular or microbial signatures improves patient outcomes. Such follow-up research will enhance the external validity of our results and advance the collective goal of refining ARDS management and prognosis.

In summary, this study leverages a rigorous multicenter design and multi-omics approaches to address critical knowledge gaps in ARDS onset and progression. By focusing on host-microbe interactions, and predictive biomarkers, it aligns with the precision medicine agenda and paves the way for more effective, personalized care in this challenging patient population.

### Trial status

The first patient was recruited on November 15, 2023, and is expected to be completed by November 2025. Current protocol version is 1.1.

## Data Availability

Deidentified research data will be made publicly available when the study is completed and published.

## Authors’ contributions

Z. Duan, D. Lian and K. Wang conceived of the original idea for the trial, have been part of the trial design and protocol writing, edited the paper, and were overall guarantors. L. Xie obtained ethical approval and has been part of the trial design as well as drafted the protocol. F. Xie, W. Xie and Z. Han have contributed to the study design and interpretation of the results and commented on the paper. All authors approved the final manuscript.

## Funding

This work was supported by Capital Health Development Research Special Project (2022-1-5091) and Beijing Natural Science Foundation Program (7222181). The authors are solely responsible for the design and conduct of this study. The funding agency has no role in the design of this trial and will not have any role during its execution, analyses, interpretation of the data, or decision to submit results.

## Declarations Competing interests

The authors declare that they have no competing interests.

## Notes

### Clinical Trial

NCT 05922826

### Funding Statement

Yes

### Author Declarations

This trial adheres to the Declaration of Helsinki and the guidelines for the quality management of drug clinical trials. Prior to enrollment, each subject or their legal representative will receive a comprehensive explanation of the study and will be required to sign the informed consent form (Supplementary materials: additional file 1). Subject data stored in the DMS will be password-protected, with access restricted exclusively to authorized users at the appropriate permission level to ensure the security of data used for statistical analysis. This study has been approved by the Ethics Committee of the PLA General Hospital (Approval Number: 2022113030901836).

